# Circulating Apolipoprotein Profiles and Their Associations with Mild Cognitive and Behavioral Impairment in Older Adults: A Cross-Sectional Study

**DOI:** 10.64898/2026.11.14.26368084

**Authors:** Arisara Amrapala, Solaphat Hemrungrojn, Natawut Nupairoj, Marco Solmi, Chavit Tunvirachaisakul, Michael Maes

## Abstract

**Background:** Mild cognitive impairment (MCI) and mild behavioral impairment (MBI) are overlapping prodromal states associated with dementia risk. Circulating apolipoproteins (Apos) may be linked to cognitive and behavioral changes, but their relationships remain unclear.

**Objective:** Examine plasma concentrations of multiple Apos in prodromal dementia.

**Methods:** This cross-sectional study included 148 Thai participants aged 60-75 years: 56 cognitively normal controls, 47 with MCI without MBI, and 45 with MBI with or without MCI. We measured ApoA1, ApoA2, ApoB, ApoC1, ApoC3, ApoD, ApoE, ApoH, and ApoJ, five lipids, and *APOE* genotype, and computed an atherogenic Apo composite (ApoB-ApoC1-ApoC3). Primary analyses used multivariable linear regression of quantitative cognitive and behavioral outcomes. Group comparisons were secondary and served to contextualize the dimensional regression findings.

**Results:** Across Montreal Cognitive Assessment (MoCA) models, 20.7-28.2% of variance was explained by combinations of ApoH, ApoC3, *APOE* ε4 carrier status, age, and male sex (inverse associations), and free cholesterol and education (positive associations). The atherogenic composite was inversely associated with MoCA total, memory index, and delayed recall scores, and with Consortium to Establish a Registry for Alzheimer’s Disease (CERAD) verbal fluency and word list recall. No significant predictors emerged for Neuropsychiatric Inventory (NPI) scores or NPI caregiver distress. All nine Apos differed significantly between groups, with levels highest in MCI, intermediate in MBI, and lowest in controls.

**Conclusions:** Circulating Apo profiles differed across controls, MCI, and MBI groups and were more consistently associated with cognitive than behavioral outcomes. ApoH and the atherogenic Apo composite warrant further investigation as candidate biomarkers of early cognitive dysfunction.

## Introduction

Global aging brings increased risk, prevalence, and public-health burden of dementia, a syndrome characterized by progressive cognitive decline and behavioral changes. As neurodegeneration involves complex interactions across multiple biological systems, current interventions cannot reliably reverse or halt the underlying pathology once dementia has manifested.^1^ With this limitation, early diagnosis and prevention are key in dementia research. Newer work has shifted towards enhancing the detection of prodromal stages of dementia, particularly mild cognitive impairment (MCI) and mild behavioral impairment (MBI).^2,3^ MCI describes an objective phase of cognitive decline that does not extensively compromise independent functioning,^4^ whilst MBI describes the later-life emergence of persistent neuropsychiatric symptoms that often accompany or precede any cognitive symptoms.^5^ Both syndromes may be associated with an elevated risk of subsequent dementia, with an annual conversion rate of approximately 10-20% from MCI^6,7^ and 14.7% from MBI,^8^ although progression estimates vary.

Vascular risk factors are among the most well-established clinical risk factors for the diagnosis and pathogenesis of all-cause dementia.^9–14^ Modifiable vascular risk factors such as diabetes and hypertension contribute to neurodegeneration, whilst vascular ageing interferes with neuronal survival.^14^ Although certain treatments or lifestyle modifications seem to effectively reduce dementia risk,^15–19^ their success is contingent on factors such as age, and the interplay of other risk factors. An estimated 45% of dementia cases may be attributed to potentially modifiable risk factors,^20,21^ highlighting the importance of early biomarker detection during midlife.

Among the biomarkers, circulating apolipoproteins (Apos) are plausible candidate markers of dementia-related risk and pathology. Apos are a group of specialized, multifunctional amphipathic proteins responsible for the stabilization and regulated transport of lipoproteins by binding to lipid surfaces, with different Apo species playing distinct biological roles.^22,23^ Their relevance to dementia is supported by reported associations with neurodegeneration and cognitive function.^24–27^ To date, previous work has explored the role of circulating Apos in cognitive impairment; however, their relationship with MBI remains largely unexplored.

Circulating ApoE has been the most extensively studied species in dementia, and its concentrations may provide information beyond *APOE* genotype. In a population study of over 76,000 individuals, lower plasma ApoE was associated with AD and all-cause dementia independent of *APOE* genotype and cardiovascular risk factors.^28^ Neuropathological work on amygdalae specimens from dementia cases has also demonstrated ApoE enrichment and colocalization with complement component C4 in amyloid plaques, including among individuals without *APOE* ε4.^29^ Longitudinally, lower plasma ApoE in MCI was associated with subsequent progression to AD, poorer cognition, and adverse amyloid, tau, and neurofilament light chain profiles.^30^

Research in other Apo species is more heterogeneous and varies by clinical population and biospecimen. Two meta-analyses reported lower circulating ApoA1 levels in AD than in controls,^27,31^ whereas an inverse relationship between ApoA1 and cognition was observed in a type 2 diabetes cohort with cognitive impairment.^32^ Higher CSF ApoB has been associated with early tau dysregulation and subsequent visuospatial decline,^33^ while higher plasma ApoC3 and ApoJ have been linked to higher baseline cognitive function or lower dementia risk in longitudinal studies.^25,34^ Multi-Apo profiling has also identified lower ApoA1, ApoA2, and ApoH, but higher ApoE, ApoJ, and ApoB/ApoA1 ratios in MCI individuals that seemed to correspond with cognitive performance, dementia risk, and brain structure changes.^26^ Despite research interests in Apos and cognitive impairment, the available body of knowledge remains inconsistent in establishing circulating Apos as reliable biomarkers of dementia risk.^11^

Direct evidence concerning circulating Apo profiles in MBI remains limited, as previous studies have focused primarily on *APOE* genotype.^35,36^ The present study therefore aimed to examine plasma concentrations of multiple Apos and conventional lipid measures in individuals with MCI without MBI and individuals with MBI with or without co-occurring MCI, compared to controls. By analyzing these profiles across participant groups, we sought to clarify whether specific Apo species or composites, along with other demographic factors, are associated with early cognitive or behavioral impairment that could potentially serve as additional candidate correlates of pre-dementia risk.

## Methods

### Participants

This cross-sectional study recruited 148 older adults aged 60-75 years from King Chulalongkorn Memorial Hospital (KCMH), the Thai Red Cross Society, and community-based senior groups in Thailand. The sample comprised 56 cognitively normal healthy controls who exhibited cognitive and behavioral performance within the normative range, 47 participants with MCI without MBI, and 45 individuals with MBI with (25) or without (20) co-occurring MCI.

Controls were included in the study if they did not meet the study criteria for MCI or MBI, had a Thai Mini-Mental State Examination (TMSE) score > 23, a Clinical Dementia Rating Sum of Boxes (CDR-SB) score of 0, and no memory deterioration as assessed by the Informant Questionnaire on Cognitive Decline in the Elderly (IQCODE). MCI participants were eligible if they had a TMSE score > 23, a CDR-SB score between 0 and 0.5, subjective memory complaints, and were diagnosed by an experienced neuropsychiatrist using the Diagnostic and Statistical Manual of Mental Disorders, Fifth Edition (DSM-5) and Petersen’s Criteria. People with MBI were included if they had a TMSE score > 23, a CDR-SB score between 0 and 0.5, a Mild Behavioral Impairment Checklist (MBI-C) score > 6.5, and were clinically diagnosed by an experienced neuropsychiatrist using the Alzheimer’s Association International Society to Advance Alzheimer’s Research and Treatment (ISTAART-AA) MBI criteria. Participants who met the criteria for both MCI and MBI were classified into the MBI cohort.

Exclusion criteria included dementia; major neurological disorders other than the conditions under study, including multiple sclerosis and stroke; lifetime and current major depressive, bipolar, psychotic, or substance-use disorders; neurodevelopmental disorders; diagnosis of medical disorders such as chronic kidney disease, chronic obstructive pulmonary disease, inflammatory bowel disease, rheumatoid arthritis, and thyroid disorders; abnormal liver and kidney blood tests; and cognitive training within the previous three months.

### Data collection

#### Participant screening

Potential participants were first informed about the study scope and procedures. Those who remained interested then completed screening assessments, namely the TMSE, CDR-SB, IQCODE, and MBI-C. For individuals with existing hospital medical records, their physicians were contacted to discuss and confirm their neuropsychiatric diagnoses. For those without prior neuropsychiatric examination or hospital records, an appointment with an experienced neuropsychiatrist was arranged to confirm eligibility before enrollment.

#### Data collection procedure

Data were collected between June 2024 and October 2025 at KCMH over one or two visits, depending on participant availability and ability. After written consent, sociodemographic, medical, medication, and supplement histories were recorded. Then, 40 mL of fasting (8 hours) blood was collected between 7 and 8 a.m., and participants were given one hour to rest. Participants then returned for neuropsychological and behavioral testing. Participants could take breaks between each assessment as needed and were compensated after protocol completion. Informants were contacted and interviewed within one week of the participant visit.

#### Blood

Following collection, fasting blood samples were immediately transported to the hospital laboratory for processing. Samples were centrifuged and separated into whole blood, serum, and plasma fractions. Then, some samples were sent to the relevant laboratories for immediate analysis, whilst the rest were stored at −80°C until thawed for further analysis.

### Measures

#### Sociodemographic, physical measurements, and history

Recorded sociodemographic variables included age, sex, marital status, number of years of education, religion, income, employment history, living conditions, and family history. Anthropometric measurements included height, weight, and waist circumference. Medical history, medication, and supplement use were also recorded.

#### Quantification of Apos and lipid profile from plasma samples

Plasma Apo concentrations were quantified using the Bio-Plex Pro^TM^ Human Apolipoprotein Assay 10-Plex Panel Kit (Catalog #12003081, Bio-Rad Laboratories, Hercules, CA, USA) on a Bio-Plex Suspension Array System (Luminex 100, Bio-Rad, Hercules, CA). The following Apo species were detected: ApoA1, ApoA2, ApoB, ApoC1, ApoC3, ApoD, ApoE, ApoH, and ApoJ. This assay uses a multiplex fluorescent immunoassay platform based on uniquely fluorescently dyed beads (xMAP technology).

High-density lipoprotein cholesterol (HDL-C), total cholesterol, and triglyceride concentrations were measured in the hospital laboratory using the Alinity C system (Abbott Laboratories, USA; Otawara-Shi, Tochigi-Ken, Japan) with enzymatic, accelerator-selective detergent, and glycerol phosphate oxidase methods, respectively. The coefficients of variation (CV) were 2.6% for HDL-C, 2.3% for total cholesterol, and 2.3% for triglycerides. Low-density lipoprotein cholesterol (LDL-C) was estimated using the Friedewald equation. Plasma free cholesterol levels were determined using the Free Cholesterol Assay Kit (Catalog #E-BC-K004- M, Elabscience, Houston, TX, USA) according to the manufacturer’s protocol.

Based on prespecified biological classifications, ApoA1, ApoE, and ApoJ were grouped as the reputedly protective composite because of their reported antioxidant and anti-inflammatory functions.^37–39^ ApoB, ApoC1, and ApoC3 were grouped as the reputedly atherogenic Apo composite due to their reported proatherogenic or inflammatory associations.^40–42^ The remaining species, ApoA2, ApoD, and ApoH, were deemed to have more context-dependent biological roles and were considered separately. The ApoB–ApoA1, atherogenic– protective Apo, LDL-C–HDL-C, and triglyceride–HDL-C indices were calculated as differences between the standardized Z-scores of their respective components.

#### APOE genotyping

Genomic DNA was extracted from peripheral blood samples using the TIANamp Genomic DNA Kit (TIANGEN Biotech, Beijing, China), according to the manufacturer’s instructions. DNA concentration and purity were assessed before downstream analysis. The target region of the *APOE* gene was amplified by polymerase chain reaction (PCR), and the PCR products were examined by agarose gel electrophoresis. Participants were subsequently classified as either having ‘No ε4 allele’ or having ‘At least one ε4 allele’ based on the identified sequence variants.

#### Montreal Cognitive Assessment (MoCA)

The MoCA is a highly sensitive, widely validated tool for detecting MCI and early AD.^43^ The test comprises 30 total points that determine the individual’s performance across eight cognitive domains: visuospatial function, executive function, memory, naming, language, attention, abstraction, and orientation. In addition to the total score, MoCA subdomain scores can help further differentiate individuals into distinct cognitive syndromes,^44^ whilst the Memory Index Score (MoCA-MIS), a recall score for the memory items, may help further discriminate between normal cognition and MCI^45^ and indicate the extent of disease progression. In this study, we used the validated Thai translation of the MoCA (MoCA-Thai).^46^

#### Neuropsychiatric Inventory (NPI)

Neuropsychiatric symptoms of dementia were comprehensively assessed using the NPI, a validated informant-based interview.^47^ The tool assesses ten behavioral and two neurovegetative areas: delusions, hallucinations, agitation/aggression, depression/dysphoria, anxiety, elation/euphoria, apathy/indifference, disinhibition, irritability/lability, aberrant motor behavior, sleep and nighttime behavior disorders, and appetite and eating disorders. For each domain, informants first indicate whether symptoms are present. For any present symptom, frequency (1-4), severity (1-3), and associated caregiver distress (0-5) levels are rated. Frequency and severity scores are then multiplied and summed to produce a total NPI score, whilst caregiver distress scores are summed to give a total caregiver distress score.

#### The Consortium to Establish a Registry for Alzheimer’s Disease (CERAD) neuropsychological battery

The CERAD neuropsychological battery comprises standardized cognitive tests designed to assess cognitive function and supports the evaluation of AD and dementia progression.^48–50^ Our study focused on three subtests: verbal fluency (VF), word list learning (WLL), and word list recall (WLR). In the VF task, participants have to name as many animals as possible in one minute. For the WLL, participants are presented with a list of words to learn across multiple trials, and after a short delay of approximately 5 minutes, are asked to recall the word list for the WLR task.

### Statistical analysis

Statistical analyses were performed using SPSS version 32 (IBM Corp., Armonk, NY, USA) and graphs were generated using MATLAB version R2026b (MathWorks, Natick, MA, USA). Continuous variables were examined for normality using histograms, Q-Q plots, and the Shapiro-Wilk test, and are presented as mean (± SD) or median (interquartile range), as appropriate. Categorical variables are presented as number (percentage). Statistical significance was set at *p* < 0.05. There were no missing data for the variables included in the primary analyses. The study population was selected to encompass a broad spectrum of cognitive and behavioral functioning, including cognitively normal individuals and participants meeting conventional criteria for MCI or MBI. Rather than treating these diagnostic categories as the principal outcomes, we focused on continuous measures of the cognitive and behavioral phenome.^51,52^ Accordingly, multivariable linear regression was the primary analytical approach, while comparisons between diagnostic groups served a secondary purpose, providing clinical context and facilitating comparisons with existing literature.

For each analyte, Z-scores were calculated using SPSS based on the entire sample. Composite values were calculated as the sum of the corresponding component Z-scores, with each component equally weighted. Z-score standardization eliminated differences in measurement units and concentration scales, ensuring each Apo or lipid contributed equally in terms of standardized variability and preventing analytes with larger absolute concentrations or variances from disproportionately influencing the composite indices. Principal component analysis (PCA) was performed on the nine Apos to determine whether a common underlying dimension could be extracted. Sampling adequacy and the suitability of the correlation matrix for PCA were assessed using the Kaiser-Meyer-Olkin (KMO) measure and Bartlett’s test of sphericity, respectively. The first principal component (PC1) was considered acceptable when the KMO value exceeded 0.70, Bartlett’s test was significant (*p* < .05), PC1 explained more than 50% of the total variance, and all nine Apos showed loadings greater than .70. One-way Analysis of Variance (ANOVA) and Kruskal-Wallis tests were used to compare continuous variables among groups, while pairwise comparisons were examined with Bonferroni-adjusted contrasts. When the homogeneity-of-variance assumption was not met, Welch’s ANOVA with Games- Howell post hoc comparisons were used. Categorical variables were compared using Pearson’s chi-square test, while individual NPI symptom frequencies were compared using two-sided Fisher-Freeman-Halton exact tests.

Apo and lipid concentration differences between groups were examined using general linear models adjusted for age, sex, BMI, and metabolic-syndrome status. Estimated marginal means, 95% confidence intervals, and omnibus tests were reported. To account for multiple testing across related biomarker outcomes, Benjamini-Hochberg false discovery rate (BH-FDR) *p*-value correction was applied separately within four prespecified families: nine individual apolipoproteins, five individual lipid measures, four derived Apo measures, and two derived lipid measures. As a sensitivity analysis, MBI participants were stratified according to the presence or absence of co-occurring MCI. Apo levels were compared between MBI-only and MBI with co-occurring MCI participants using general linear models with age and BMI as covariates and sex and metabolic-syndrome status as fixed factors. BH-FDR correction was applied across the nine apolipoprotein comparisons. Multivariable linear regression was used to examine associations with cognitive and global behavioral outcomes. For each outcome, one model evaluated individual Apo species together with lipid, demographic, and *APOE* variables, and an alternative model replaced individual Apo candidates with the prespecified composites (protective and atherogenic composites) together with the remaining candidate predictors.

The multivariable regression analyses constituted the primary outcome analyses, focusing on quantitative cognitive and behavioral measures rather than categorical diagnoses. Comparison among diagnostic groups was presented as supplementary descriptive analyses to facilitate clinical interpretation, particularly for readers less familiar with dimensional regression approaches. Standardized regression coefficients (β), t-statistics, *p*-values, and model-fit statistics were reported. Assumptions of the multivariable regression models were evaluated using regression diagnostics. Multicollinearity was assessed using tolerance and variance inflation factor (VIF) values. Normality of residuals was examined using residual histograms and normal probability (P-P) plots of standardized residuals, while homoscedasticity was visually assessed using plots of standardized residuals against standardized predicted values. An a priori power analysis for multivariable linear regression, assuming seven predictors, an anticipated explained variance of 15% (R^2^ = .15), and α = .05, indicated that a minimum of 89 participants were required to achieve 80% power. Given the final sample of 148 participants, the estimated achieved post-hoc power was approximately 97.3%.

## Results

### Participant demographic and clinical characteristics

The study included 148 participants (**Table 1**). The groups did not differ significantly in age, sex distribution, BMI, metabolic-syndrome prevalence, or *APOE* ε4-carrier status. Years of education and IQCODE scores differed significantly across groups, with adjusted pairwise comparisons indicating differences between controls and MCI, but not between MBI and either the controls or MCI. Additionally, significant group differences were observed in TMSE, CDR-SB, and MBI-C scores, consistent with the clinical characteristics used to define the study group: MCI participants had the lowest TMSE scores and highest CDR-SB scores, whereas MBI participants had the highest MBI-C scores.

**Table 1.** Demographic and Clinical Characteristics in Controls, Mild Cognitive Impairment (MCI), and Mild Behavioral Impairment (MBI)

|  | **All<br>(N = 148)** | **Controls<br>(N = 56)** | **MCI<br>(N = 47)** | **MBI<br>(N = 45)** | **Test Statistic** | **df** | ***p*-value** |
|---|---|---|---|---|---|---|---|
| **Demographic Characteristics** |  |  |  |  |  |  |  |
| **Age**, years | 67.2 ± 4.11 | 66.5 ± 3.98 | 68.4 ± 4.35 | 66.9 ± 3.83 | *F* = 2.94 | 2, 145 | .056 |
| **Sex**, n (%) |  |  |  |  | χ<sup>2</sup> = 1.06 | 2 | .589 |
| Males | 44 (29.7) | 14 (25.0) | 16 (34.0) | 14 (31.1) |  |  |  |
| Females | 104 (70.3) | 42 (75.0) | 31 (66.0) | 31 (68.9) |  |  |  |
| **BMI**, kg/m<sup>2</sup> | 23.5 ± 3.57 | 23.2 ± 3.35 | 23.9 ± 3.60 | 23.7 ± 3.85 | *F* = 0.555 | 2, 145 | .575 |
| **Years of Education** | 16.0 [14.0-16.0] | 16.0 [16.0-18.0]<sup>a</sup> | 16.0 [11.0-16.0]<sup>b</sup> | 16.0 [14.5-18.0]<sup>ab</sup> | *H* = 8.68 | 2 | .013* |
| **Clinical Characteristics** |  |  |  |  |  |  |  |
| **Metabolic Syndrome**, n (%) | 23 (15.5) | 7 (12.5) | 11 (23.4) | 5 (11.1) | χ<sup>2</sup> = 3.28 | 2 | .194 |
| **TMSE Score** | 29.0 [27.0-30.0] | 29.0 [28.0-30.0]<sup>a</sup> | 28.0 [27.0-29.0]<sup>b</sup> | 29.0 [27.0-30.0]<sup>ab</sup> | *H* = 12.9 | 2 | **.002**\* |
| **CDR-SB Score** | 0.00 [0.00-0.50] | 0.00 [0.00-0.00]<sup>a</sup> | 0.50 [0.50-0.50]<sup>b</sup> | 0.50 [0.00-0.50]<sup>c</sup> | *H* = 103 | 2 | **<.001**\*\* |
| **MBI-C Score** | 2.00 [0.00-8.00] | 0.00 [0.00-2.00]<sup>a</sup> | 1.00 [0.00-3.00]<sup>a</sup> | 11.0 [8.25-18.8]<sup>b</sup> | *H* = 97.8 | 2 | **<.001**\*\* |
| **IQCODE Score** | 3.13 [3.00-3.31] | 3.06 [3.00-3.19]<sup>a</sup> | 3.22 [3.05-3.38]<sup>b</sup> | 3.13 [3.00-3.31]<sup>ab</sup> | *H* = 8.77 | 2 | .012* |
| ***APOE* genotype**, n (%) |  |  |  |  | χ<sup>2</sup> = 2.89 | 2 | .235 |
| No ε4 allele | 119 (80.4) | 49 (87.5) | 36 (76.6) | 34 (75.6) |  |  |  |
| ≥ 1 ε4 allele | 29 (19.6) | 7 (12.5) | 11 (23.4) | 11 (24.4) |  |  |  |

Normally distributed continuous variables are presented as *mean (±SD)*; *median* [Q1-Q3] for skewed or ordinal continuous variables, and *n* (%) for categorical variables. Between-group comparisons were performed using one-way analysis of variance (ANOVA) for approximately normally distributed continuous variables, Kruskal-Wallis tests for skewed or ordinal continuous variables, and Pearson’s chi-square test for categorical variables, as appropriate. For significant Kruskal-Wallis tests, pairwise comparisons were performed with Bonferroni adjustment. Within each row, groups sharing a superscript letter did not differ significantly, whereas groups with no superscript letter in common differed at Bonferroni-adjusted *p* < .05.
*F*, one-way ANOVA statistic; *H*, Kruskal–Wallis statistic; χ², Pearson chi-square statistic; \**p* < .05; \*\**p* < .001
BMI: Body Mass Index; TMSE: Thai Mini-Mental State Examination; CDR-SB: Clinical Dementia Rating Sum of Boxes; MBI-C: Mild Behavioral Impairment Checklist; IQCODE: Informant Questionnaire on Cognitive Decline in the Elderly; APOE: Apolipoprotein E.

### Cognitive and behavioral characteristics

MCI participants generally demonstrated poorer cognitive performance, particularly on memory-sensitive outcomes (**Table 2**). The MCI cohort had a significantly lower MoCA total score than both the MBI and control groups. Significant group differences were also observed for the MoCA-MIS, Visuospatial/Executive, Attention, Language, Abstraction, and Delayed Recall subdomains. No significant group differences were found in Naming and Orientation. The largest group differences were observed for the MoCA total score, delayed recall subdomain, and MoCA-MIS. For the CERAD outcomes, MCI participants showed poorer performance overall. Significantly lower WLL and WLR scores were obtained in the MCI group than in controls and MBI participants, whereas for VF, significant differences were found between MCI participants and controls, but not with MBI.

**Table 2.** Cognitive and Behavioral Characteristics in Controls, Mild Cognitive Impairment (MCI), and Mild Behavioral Impairment (MBI)

|  | **Controls<br>(N = 56)** | **MCI<br>(N = 47)** | **MBI<br>(N = 45)** | **Test<br>Statistic** | **dF** | ***p*-value** |
|---|---|---|---|---|---|---|
| **Cognition: MoCA Scores and Subdomains** |  |  |  |  |  |  |
| **MoCA Total Score** (/30) | 27.0 [26.0-29.0]<sup>a</sup> | 22.0 [20.0-24.0]<sup>c</sup> | 25.0 [23.0-27.0]<sup>b</sup> | *H* = 74.4 | 2 | **<.001**\*\* |
| **MoCA-MIS** (/15) | 15.0 [13.0-15.0]<sup>a</sup> | 8.50 [6.75-10.0]<sup>c</sup> | 12.0 [9.25-14.0]<sup>b</sup> | *H* = 64.9 | 2 | **<.001**\*\* |
| **Visuospatial/Executive Function** (/5) | 5.00 [4.00-5.00]<sup>a</sup> | 4.00 [3.00-5.00]<sup>b</sup> | 5.00 [4.00-5.00]<sup>a</sup> | *H* = 17.4 | 2 | **<.001**\*\* |
| **Naming** (/3) | 3.00 [3.00-3.00] | 3.00 [3.00-3.00] | 3.00 [3.00-3.00] | *H* = 4.72 | 2 | .094 |
| **Attention** (/6) | 6.00 [5.00-6.00]<sup>a</sup> | 6.00 [5.00-6.00]<sup>ab</sup> | 5.50 [5.00-6.00]<sup>b</sup> | *H* = 8.28 | 2 | **.016**\* |
| **Language** (/3) | 2.00 [2.00-3.00]<sup>a</sup> | 2.00 [1.00-2.00]<sup>b</sup> | 2.00 [1.00-2.00]<sup>b</sup> | *H* = 12.6 | 2 | **.002**\* |
| **Abstraction** (/2) | 2.00 [1.00-2.00]<sup>a</sup> | 1.00 [1.00-2.00]<sup>b</sup> | 2.00 [1.00-2.00]<sup>ab</sup> | *H* = 10.0 | 2 | **.007**\* |
| **Delayed Recall** (/5) | 5.00 [4.00-5.00]<sup>a</sup> | 1.00 [0.00-2.00]<sup>c</sup> | 3.00 [2.00-4.00]<sup>b</sup> | *H* = 66.9 | 2 | **<.001**\*\* |
| **Orientation** (/6) | 6.00 [6.00-6.00] | 6.00 [6.00-6.00] | 6.00 [6.00-6.00] | *H* = 4.22 | 2 | .121 |
| **Cognition: CERAD Subtests** |  |  |  |  |  |  |
| **Verbal Fluency Score** (/24) | 24.0 [21.0-24.0]<sup>a</sup> | 22.5 [18.5-24.0]<sup>b</sup> | 24.0 [19.3-24.0]<sup>ab</sup> | *H* = 6.53 | 2 | **.038**\* |
| **Word List Learning Total Score** (/30) | 23.7 ± 2.71<sup>a</sup> | 20.2 ± 3.67<sup>b</sup> | 22.3 ± 3.39<sup>a</sup> | Welch *F* = 15.2 | 2, 89.9 | **<.001**\*\* |
| **Word List Recall Score** (/10) | 8.00 [7.00-9.00]<sup>a</sup> | 6.00 [4.00-7.00]<sup>b</sup> | 7.00 [6.00-8.00]<sup>a</sup> | *H* = 22.6 | 2 | **<.001**\*\* |
| **Behavior: NPI Scores and Presence of Individual Symptoms** |  |  |  |  |  |  |
| **Total NPI Score** | 0.00 [0.00-0.00]<sup>a</sup> | 0.00 [0.00-1.00]<sup>a</sup> | 1.50 [0.00-4.75]<sup>b</sup> | *H* = 21.3 | 2 | **<.001**\*\* |
| **Total Caregiver Distress Score** | 0.00 [0.00-0.00]<sup>a</sup> | 0.00 [0.00-0.00]<sup>a</sup> | 0.500 [0.00-3.00]<sup>b</sup> | *H* = 24.3 | 2 | **<.001**\*\* |
| **Delusion** | 1 (1.79) | 0 (0) | 2 (4.44) | - | - | .394 |
| **Hallucination** | 1 (1.79) | 0 (0) | 0 (0) | - | - | 1.00 |
| **Agitation** | 4 (7.14) | 6 (12.8) | 11 (24.4) | - | - | .055 |
| **Depression/Dysphoria** | 3 (5.36) | 1 (2.13) | 4 (8.89) | - | - | .390 |
| **Anxiety** | 1 (1.79) | 2 (4.26) | 9 (20.0) | - | - | **.003**\* |
| **Euphoria/Elation** | 1 (1.79) | 1 (2.13) | 1 (2.22) | - | - | 1.00 |
| **Apathy/Indifference** | 4 (7.14) | 3 (6.38) | 7 (15.6) | - | - | .279 |
| **Disinhibition** | 2 (3.57) | 2 (4.26) | 11 (24.4) | - | - | **.001**\* |
| **Irritability/Lability** | 1 (1.79) | 1 (2.13) | 14 (31.1) | - | - | **<.001**\*\* |
| **Aberrant Motor Activity** | 0 (0) | 3 (6.38) | 6 (13.3) | - | - | **.010**\* |
| **Nighttime Behavior** | 5 (8.93) | 5 (10.6) | 4 (8.89) | - | - | 1.00 |
| **Appetite/Eating Changes** | 1 (1.79) | 1 (2.13) | 2 (4.44) | - | - | .688 |

Normally distributed continuous variables are presented as *mean (±SD)*; median [Q1-Q3] for skewed or ordinal continuous variables, and n (%) for categorical variables. Between-group comparisons were performed using Kruskal-Wallis tests with Bonferroni-adjusted pairwise comparisons for continuous outcomes, except for CERAD WLL, which was assessed using Welch’s one-way analysis of variance (ANOVA) with Games-Howell post hoc comparisons because the homogeneity-of-variance assumption was not met. Presence of individual NPI symptoms was compared using two-sided Fisher-Freeman-Halton exact tests. Within each continuous-outcome row, groups sharing a superscript letter did not differ significantly, whereas groups with no superscript letter in common differed at adjusted *p* < .05.
*H*, Kruskal–Wallis statistic; χ², Pearson chi-square statistic; \**p* < .05; \*\**p* < .001
MoCA: Montreal Cognitive Assessment; MIS: Memory Index Score; CERAD: The Consortium to Establish a Registry for Alzheimer’s Disease; NPI: Neuropsychiatric Inventory.

Behaviorally, the MBI cohort had significantly higher global NPI total score and caregiver distress than both the controls and MCI cohorts, whereas controls and MCI did not differ significantly on either measure (Table 2). Significant differences in individual symptom prevalence were observed for Anxiety, Disinhibition, Irritability/Lability, and Aberrant Motor Activity, with the MBI cohort having the highest prevalence. The remaining symptoms did not differ significantly.

### Apo and lipid plasma concentrations

After adjustment for age, sex, BMI, and metabolic-syndrome status, all individual Apo species showed significant diagnostic-group effects. Adjusted means were highest in MCI participants, intermediate in MBI, and lowest in controls (**Figure 1A**; Supplementary Table S1). However, specific pairwise differences varied among Apos (Supplementary Table S2). The protective and atherogenic Apo composites also differed significantly across diagnostic groups, but when the atherogenic-protective Apo composite difference was considered, no significant differences across groups were found. For the ApoB-ApoA1 difference, there was a smaller omnibus group effect, although none of the Bonferroni-adjusted pairwise comparisons reached significance.

**Figure 1.** Plasma Apolipoprotein and Lipid Profiles by Diagnostic Group. (A) Means of individual plasma apolipoprotein species and (B) lipid means in controls, MCI, and MBI participants. Bars indicate adjusted marginal mean Z-scores, and error bars indicate ± 1 standard error. Models were adjusted for age and body mass index, with sex and metabolic-syndrome status included as fixed factors.

Lipid profile patterns were less uniform: total cholesterol and triglycerides were highest in MCI, HDL-C was lowest in controls, and LDL-C was lowest in MBI; however, none of the five individual lipid measures showed a significant adjusted diagnostic-group effect (Figure 1B; Supplementary Table S1). The LDL-C-HDL-C difference showed a borderline omnibus effect before multiple-testing correction but did not remain significant after BH-FDR correction. The Triglyceride-HDL-C difference was also nonsignificant.

After BH-FDR correction, significant diagnostic group effects remained for all nine Apos, the protective and atherogenic Apo composites, and the ApoB-ApoA1 difference. No individual lipid measure or derived lipid difference remained significant after BH-FDR correction.

PCA identified a single dominant Apo component explaining 75.79% of the total variance, with all nine Apos showing strong positive loadings (.814-.939), KMO = 0.919, Bartlett’s test significant (χ^2^ = 1412.58, df = 36, *p* < .001), labeled as PC-Apo score (Supplementary Table S1). This component represents a common dimension of circulating Apo abundance, whereby higher component scores reflect coordinated elevations across all nine Apos. Thus, the PC-Apo score captures shared variation across Apos with diverse biological functions rather than a specific atherogenic or protective profile. PC-Apo scores differed significantly between groups, with higher scores in both MCI and MBI than in controls (both *p* < .001). Although scores were higher in MCI than in MBI, this difference was not statistically significant (*p* = .350).

In sensitivity analyses restricted to the MBI cohort, the Apo levels were consistently higher among participants with co-occurring MCI than among those without MCI. Nominal subgroup differences were observed for ApoA2, ApoC1, ApoC3, and ApoJ; however, none remained significant after BH-FDR correction across the nine Apos.

No significant differences in individual Apo levels were found between *APOE* genotype groups, and *APOE* ε4-carrier status prevalence did not differ significantly across diagnostic groups (**Figure 2**).

**Figure 2.** Apolipoprotein Profiles and APOE ε4-Carrier Prevalence by APOE genotype and Diagnostic Group. (A) Mean plasma apolipoprotein species in participants without an *APOE* ε4 allele and those carrying at least one ε4 allele. Bars represent group means and error bars indicate ± 1 standard error. Models were adjusted for age and body mass index, with sex and metabolic- syndrome status included as fixed factors. No significant differences were observed between non-carriers and carriers (all *p* > .05). (B) Prevalence of *APOE* ε4 carriers across controls, mild cognitive impairment (MCI), and mild behavioral impairment (MBI) cohorts. Bars represent observed proportions and error bars indicate 95% Wilson confidence intervals. Carrier prevalence did not significantly differ across diagnostic groups (*p* = .235).

### The relationship between Apos, lipids, cognition, and behavior

Multiple regression analysis generated twelve final models. In models containing individual Apo species (**Table 3**), ApoH was inversely associated with MoCA total score (**Figure 3A**), MoCA-MIS, CERAD WLL, and CERAD WLR. ApoC3 was inversely associated with MoCA delayed recall (Figure 3C) and CERAD VF. Higher free cholesterol was associated with better MoCA total score, MIS (Figure 3B), and delayed recall performance, while LDL-C was positively associated with VF. *R*^2^ values ranged from 0.116 for WLL to 0.282 for MoCA total score. Cognitive measures from the MoCA and CERAD were associated with at least one Apo species (Table 3).

**Figure 3.** Partial Regression Plots for Selected Associations Between Cognitive Outcomes and Biochemical Predictors in the Multivariable Linear Regression Models. Panels show the relationship between (A) ApoH and MoCA total score, (B) Free cholesterol and MoCA-memory index score, (C) ApoC3 and MoCA delayed recall subdomain Score, and (D) Atherogenic Apo composite and MoCA total score. Each plot represents the relationship between the indicated predictor and outcome after accounting for the other variables included in the corresponding regression model.

**Table 3.** Multiple Regression of the Cognitive Measures on Apolipoprotein and Lipid Variables.

| **Dependent Variables** | **Explanatory Variables** | **Coefficients of Input Variables** |  |  | **Model Statistics** |  |  |  |
|---|---|---|---|---|---|---|---|---|
|  |  | **β** | **t** | ***p*** | **Adjusted R<sup>2</sup>** | **dF** | **F** | ***p*** |
| **MoCA Total Score** | **Model #1** |  |  |  | 0.282 | 3, 144 | 20.3 | <.001** |
|  | Years of Education | 0.298 | 4.23 | <.001 |  |  |  |  |
|  | Free Cholesterol | 0.241 | 3.44 | <.001 |  |  |  |  |
|  | ApoH | -0.345 | -4.91 | <.001 |  |  |  |  |
|  | **Model #2** |  |  |  | 0.254 | 3, 144 | 17.7 | <.001** |
|  | Years of Education | 0.296 | 4.1 | <.001 |  |  |  |  |
|  | Atherogenic Apos | -0.304 | -4.22 | <.001 |  |  |  |  |
|  | Free Cholesterol | 0.236 | 3.31 | .001 |  |  |  |  |
| **MoCA MIS** | **Model #3** |  |  |  | 0.239 | 4, 143 | 12.5 | <.001** |
|  | ApoH | -0.334 | -4.6 | <.001 |  |  |  |  |
|  | Free Cholesterol | 0.245 | 3.38 | <.001 |  |  |  |  |
|  | *APOE* Genotype | -0.193 | -2.66 | .009 |  |  |  |  |
|  | Age | -0.153 | -2.12 | .036 |  |  |  |  |
|  | **Model #4** |  |  |  | 0.232 | 5, 142 | 9.89 | <.001** |
|  | Atherogenic Apos | -0.294 | -4.03 | <.001 |  |  |  |  |
|  | Free Cholesterol | 0.252 | 3.46 | <.001 |  |  |  |  |
|  | *APOE* Genotype | -0.217 | -2.98 | .003 |  |  |  |  |
|  | Age | -0.194 | -2.66 | .009 |  |  |  |  |
|  | Sex | -0.189 | -2.56 | .011 |  |  |  |  |
| **MoCA Delayed Recall Subdomain Score** | **Model #5** |  |  |  | 0.236 | 5, 142 | 10.1 | <.001** |
|  | ApoC3 | -0.361 | -4.92 | <.001 |  |  |  |  |
|  | Free Cholesterol | 0.208 | 2.86 | .005 |  |  |  |  |
|  | *APOE* Genotype | -0.2 | -2.75 | .007 |  |  |  |  |
|  | Sex | -0.207 | -2.81 | .006 |  |  |  |  |
|  | Age | -0.158 | -2.16 | .033 |  |  |  |  |
|  | **Model #6** |  |  |  | 0.207 | 5, 142 | 8.65 | <.001** |
|  | Atherogenic Apos | -0.315 | -4.25 | <.001 |  |  |  |  |
|  | Free Cholesterol | 0.228 | 3.07 | .003 |  |  |  |  |
|  | *APOE* Genotype | -0.172 | -2.32 | .022 |  |  |  |  |
|  | Sex | -0.196 | -2.61 | .01 |  |  |  |  |
|  | Age | -0.175 | -2.36 | .02 |  |  |  |  |
| **CERAD Verbal Fluency Score** | **Model #7** |  |  |  | 0.137 | 4, 143 | 6.85 | <.001** |
|  | Age | -0.202 | -2.62 | .010 |  |  |  |  |
|  | ApoC3 | -0.234 | -3.03 | .003 |  |  |  |  |
|  | LDL-C | 0.191 | 2.48 | .014 |  |  |  |  |
|  | *APOE* Genotype | -0.160 | -2.08 | .040 |  |  |  |  |
|  | **Model #8** |  |  |  | 0.12 | 4, 143 | 6.01 | <.001** |
|  | Age | -0.182 | -2.27 | .025 |  |  |  |  |
|  | Years of Education | 0.136 | 1.68 | .095 |  |  |  |  |
|  | LDL-C | 0.201 | 2.52 | .013 |  |  |  |  |
|  | Atherogenic Apos | -0.188 | -2.34 | .021 |  |  |  |  |
| **CERAD Word List Learning Total Score** | **Model #9** |  |  |  | 0.116 | 3, 144 | 7.42 | <.001** |
|  | ApoH | -0.210 | -2.68 | .008 |  |  |  |  |
|  | Years of Education | 0.174 | 2.16 | .032 |  |  |  |  |
|  | Age | -0.169 | -2.1 | .038 |  |  |  |  |
|  | **Model #10** |  |  |  | 0.099 | 3, 144 | 6.37 | <.001** |
|  | Years of Education | 0.19 | 2.34 | .02 |  |  |  |  |
|  | Age | -0.209 | -2.56 | .011 |  |  |  |  |
|  | Sex | -0.164 | -2.08 | .039 |  |  |  |  |
| **CERAD Word List Recall Score** | **Model #11** |  |  |  | 0.13 | 2, 145 | 11.9 | <.001** |
|  | Age | -0.284 | -3.68 | <.001 |  |  |  |  |
|  | ApoH | -0.218 | -2.82 | .006 |  |  |  |  |
|  | **Model #12** |  |  |  | 0.108 | 2, 145 | 9.86 | <.001** |
|  | Age | -0.297 | -3.8 | <.001 |  |  |  |  |
|  | Atherogenic Apos | -0.159 | -2.04 | .043 |  |  |  |  |

\**p*<0.05; \*\**p*<0.001
MoCA: Montreal Cognitive Assessment; MIS: Memory Index Score; CERAD: The Consortium to Establish a Registry for Alzheimer’s Disease

In the alternative models, the atherogenic Apo composite was retained as a variable inversely associated with the MoCA outcomes and CERAD VF and WLR, but not WLL (example association illustrated in Figure 3D). The protective composite was not retained in any model. All alternative models had slightly lower *R*^2^ values than their corresponding individual Apo models. No candidate variable met the retention criterion for the global NPI or caregiver-distress outcomes. Individual symptom domains were not modeled due to low frequencies.

## Discussion

### Principal findings

The first major finding is that significant differences between diagnostic groups were observed for all nine individual Apos and the Apo composites, with all differences remaining significant after BH-FDR *p*-correction. In contrast, no individual lipid or derived lipid measure remained significant after BH-FDR correction. Moreover, PCA of the nine Apos yielded a single principal component (PC), with higher PC-Apo scores in both MCI and MBI groups than in controls. This finding suggests that a coordinated increase in circulating Apos may be a shared biological feature of MCI and MBI, potentially reflecting common underlying mechanisms regulating Apo metabolism. In the multivariable models, ApoH, ApoC3, and the atherogenic Apo composite were inversely associated with selected cognitive outcomes, whilst no candidate predictors were retained for the global neuropsychiatric symptom or caregiver distress measures. These findings indicate that circulating Apo profiles may be more consistently associated with cognitive performance than with global behavioral burden, though this does not exclude possible associations with individual symptoms or changes over time.

### Distinct and overlapping cognitive and behavioral phenotypes in MCI and MBI

Although the primary outcome variables in this study were continuous measures of cognitive and behavioral functioning, rather than the categorical diagnoses of MCI and MBI, it is notable that both MCI and MBI displayed distinct, but partially overlapping clinical profiles. MCI was characterized predominantly by poorer cognitive performance, particularly in learning and delayed recall, concurrently with certain behavioral symptoms, whereas MBI was characterized by greater neuropsychiatric burden and caregiver distress, alongside accompanying cognitive deficits. However, unlike cognitive deficits that may more directly reflect underlying cognitive dysfunction, behavioral symptoms may also be influenced by psychosocial circumstances and environmental factors,^53^ and in some cases, may be more closely related to these contextual influences than to the degree of cognitive impairment. Although the syndromes can coexist,^54^ they are generally considered distinguishable but related manifestations of early clinical impairment,^3,55,56^ reflecting different dimensions of cognitive and behavioral change in older adults. This distinction may also be reflected in their underlying biological correlates, as cognitive and behavioral abnormalities may have partly distinct biological associations early in the disease course,^56,57^ with increasing overlap as impairment progresses.^58^

### Apos and other metabolic correlates of behavioral impairment

We found that MCI and MBI share increased circulating Apo levels, although these alterations were associated with cognitive dysfunction rather than behavioral symptom severity or caregiver distress. Direct evidence relating circulating Apos to MBI is particularly sparse. Available findings suggest that Apo-behavior associations may vary according to Apo species, symptom domain, and follow-up interval. In a longitudinal study with cognitive impairment, higher ApoA1 was associated with a lower likelihood of apathy, higher ApoA2 was related to higher prevalence of apathy, higher ApoB and ApoC1 predicted subsequent neuropsychiatric symptoms at two years, higher ApoE was related to fewer symptoms at two years, higher ApoH was associated with anxiety, and higher ApoJ was related to higher prevalence of neuropsychiatric symptoms at one year.^59^ This heterogeneous pattern suggests that individual behavioral domains and longitudinal changes may be more informative than a single global symptom score. Evidence concerning *APOE* genotype is similarly mixed. Although one study observed that carrying an *APOE* ε4 allele increased MBI risk,^60^ other studies suggest that *APOE* ε4 may be more related to specific domains, including affective dysregulation,^35^ late-life depression,^61^ and anxiety.^62^ Associations with other symptoms remain inconsistent or insufficiently studied.^63,64^

Literature regarding lipid, metabolic, and demographic correlates of MBI is also limited and heterogeneous. Higher serum triglycerides have been reported in MBI,^65^ while dysregulation in insulin and homocysteine^66^ has also been linked with MBI-related outcomes. Correlations have been found between fasting glucose and the social inappropriateness domain of the MBI-C.^67^ Indirect evidence from related psychiatric and dementia populations has linked lipid alterations to major depressive disorder and anxiety disorder;^68^ fasting glucose to depression, anxiety, and poorer sleep quality;^67^ and higher triglyceride concentrations to greater neuropsychiatric burden and psychosis.^69^ Demographic findings are likewise inconsistent. An exhaustive systematic review found that MBI individuals often had fewer years of education and that some studies identified associations with older age or male sex; however, several studies reported no significant demographic associations.^70^ Collectively, findings from the literature suggest that MBI correlates may depend on symptom domain, study population, disease stage, and analytical approach. Future studies should further examine individual MBI-C domains and NPI symptoms longitudinally and evaluate additional candidate biomarkers of behavioral impairment.^71^

### Coordinated alterations in circulating Apo profiles in MCI and MBI

Across groups, all nine Apo species and both Apo composites showed the highest values in MCI, intermediate in the MBI group, and the lowest values in controls. The coordinated pattern suggests that MCI was accompanied by a broad alteration in circulating Apos rather than an isolated change in a single species. Direct comparison with previous research is limited because multi-Apo profiles have not, to our knowledge, been examined across the same groups as this work. The closest comparable study reported lower ApoA1, ApoA2, and ApoH, and higher levels of ApoE, ApoJ, and ApoB/ApoA1 ratios in MCI than in controls,^26^ contrasting with the broadly higher concentrations observed here. Indirect findings from related behavioral phenotypes have likewise shown heterogeneous changes in circulating ApoA and ApoB in depression,^72^ ApoJ in relation to agitation/aggression,^73^ ApoD in schizophrenia,^74^ ApoA1 and ApoB before depression-, anxiety-, or stress-related diagnoses,^75^ and ApoB in relation to short sleep duration.^76^

### ApoH and ApoC3 as correlates of cognitive impairment: potential metabolic and vascular mechanisms

ApoH was the most consistently associated analyte, showing inverse associations with global cognition, memory indexing, verbal learning, and delayed recall, independent of *APOE* ε4 genotype. Previous studies have reported associations in the opposite direction, with positive relationships between ApoH and global cognitive, memory, or visuospatial performance.^26,77^ Differences in population characteristics, cognitive measures, metabolic status, and laboratory procedures may contribute to this inconsistency.

Apart from ApoH, ApoC3 showed a more selective pattern, with negative associations with MoCA delayed recall and CERAD verbal fluency. As these outcomes involve partly different cognitive processes, these results do not establish specificity for a single cognitive domain. These findings also differ from earlier work linking higher ApoC3 concentrations to better cognitive performance,^77,78^ particularly in the memory and visuospatial domains, slower cognitive decline,^79^ and slower CSF amyloid-β42 reduction.^80^ On a broader level, higher concentrations of ApoH and ApoC3 have been suggested to indicate metabolic syndrome and increased cardiovascular risk^81,82^ due to associations with higher triglyceride levels. Therefore, the inverse associations between ApoH, ApoC3, and cognitive performance in this research may reflect shared metabolic and vascular dysregulation mechanisms that could affect cognition. Other species, including ApoJ^83^ and ApoA1,^84^ have also been proposed as candidate correlates of neurocognitive decline, although they were not retained in the present models.

### Atherogenic and protective Apo composites: associations with cognitive performance

We found that, against a background of generally elevated circulating Apo concentrations, higher levels of proatherogenic and proinflammatory Apos (composite based on ApoB, ApoC1, and ApoC3) were particularly associated with poorer cognitive performance. Thus, the atherogenic Apo composite may be a potential correlate of general cognitive impairment, though further examination is required to determine the incremental value of the composite.

Regarding lipid profile, higher free cholesterol was associated with all three modeled MoCA outcomes. A previous study reported lower free cholesterol in peripheral blood mononuclear cells in MCI.^85^ The wider lipid literature is similarly context-dependent.^20,86–92^

### Divergent associations of lipids and Apos with cognitive performance

The generalized increase in circulating Apos in MCI and MBI may reflect coordinated alterations in hepatic lipoprotein synthesis, secretion, and clearance, potentially associated with metabolic and vascular dysregulation.^93^ Alternatively, the parallel increases across Apo species may reflect changes in circulating lipoprotein particle abundance or turnover rather than independent upregulation of individual Apos.^94^ The positive association between plasma free cholesterol and cognitive performance, contrasting with the inverse associations observed for ApoH, ApoC3, and the atherogenic Apo composite, may reflect distinct aspects of lipid homeostasis. Unesterified cholesterol is essential for membrane integrity, myelination, and synaptic formation, including astrocyte-mediated cholesterol delivery to neurons.^95^ However, cerebral cholesterol metabolism is largely independent of circulating cholesterol because of the blood-brain barrier.^96^ Consequently, higher plasma free cholesterol may reflect more favorable systemic metabolic or nutritional conditions rather than directly increased cerebral cholesterol availability. Conversely, ApoC3 impairs triglyceride-rich lipoprotein clearance and promotes atherogenic remnant accumulation and vascular inflammation.^42,97^ ApoB and ApoC1 participate in lipoprotein transport and remodeling,^23^ whereas ApoH has additional phospholipid-binding and immune-coagulation functions.^98^ Thus, the divergent associations may reflect differences between systemic cholesterol status and lipoprotein-associated metabolic and vascular dysfunction, although the mechanisms underlying the observed cognitive associations remain to be established.

### Limitations and future studies

Several limitations should be addressed. First, the cross-sectional design precludes insight into temporal sequence, causality, and prognostic value. Second, this Thai cohort was predominantly female and relatively highly educated, which may limit generalizability. The MBI group also included participants with and without co-occurring MCI, whereas the MCI group excluded MBI. Diagnostic classifications of MCI and MBI represent operationally defined, partly overlapping clinical constructs with potentially limited reliability and substantial within-group heterogeneity. Therefore, our regression analyses deliberately focused on quantitative cognitive (MoCA and CERAD) and behavioral (NPI) measures rather than categorical diagnosis, thereby preserving dimensional information and avoiding potentially artificial diagnostic boundaries.

Third, the low frequency of several individual neuropsychiatric symptoms may limit domain-specific behavioral modeling.

Fourth, the multivariable analyses were conducted within the original sample; therefore, the reproducibility and generalizability of the findings require validation in independent cohorts across different countries and cultural settings.

Future studies should use longitudinal cohorts with repeated Apo measurements that are examined alongside established neurodegenerative biomarkers, metabolic and vascular measures, and neuroimaging. Further internal validation and external replication are needed to evaluate whether ApoH, ApoC3, and the atherogenic Apo composite provide reproducible information about cognitive performance beyond established demographic factors, *APOE* genotype, and conventional lipid measures. Emerging novel analytes such as ApoM may also extend the current panel and give rise to different patterns of association, as higher plasma ApoM has been reported in AD than in controls after adjustment for established risk factors.^99^

### Conclusion

In this cross-sectional case-control study of Thai older adults, MCI and MBI were characterized by distinct, but partially overlapping, cognitive and behavioral profiles. Adjusted circulating concentrations of all measured Apo species showed a consistent pattern of highest values in MCI, intermediate in the heterogeneous MBI group, and lowest values in controls, with significant diagnostic-group effects remaining after BH-FDR correction. ApoH, ApoC3, and the atherogenic Apo composite were inversely associated with certain cognitive measures. No candidate predictors were retained for global neuropsychiatric symptom or caregiver distress regression models. These exploratory findings suggest that Apo profiles may be more consistently related to cognitive performance than behavioral severity; however, further studies are required to validate these findings before diagnostic or prognostic utility of the notable Apos can be inferred.

## Acknowledgements

We would like to express our deepest gratitude to the participants and guardians who made this research possible. We also sincerely thank the hospital staff, nurses, laboratory staff, and administrators from the Chula Neuroscience Center; Cognitive Fitness Center; Center for Medical Diagnostic Laboratories; Excellence Center in Diabetes, Hormone, and Metabolism; Division of Immunology at the Department of Microbiology; and Central Laboratory at the Department of Laboratory Medicine, all of which are part of the Faculty of Medicine, Chulalongkorn University/KCMH, Thai Red Cross Society, for their support and cooperation.

## Author contributions

**Arisara Amrapala**: Conceptualization, Methodology, Software, Validation, Formal Analysis, Investigation, Resources, Data Curation, Writing - Original Draft, Writing - Review & Editing, Visualization, Supervision, Project Administration, Funding Acquisition. **Solaphat Hemrungrojn**: Validation, Resources, Supervision, Funding Acquisition. **Natawut Nupairoj**: Resources, Funding Acquisition. **Marco Solmi:** Writing – Review & Editing, Supervision. **Chavit Tunvirachaisakul**: Conceptualization, Methodology, Validation, Resources, Supervision, Funding Acquisition. **Michael Maes**: Conceptualization, Methodology, Software, Validation, Formal Analysis, Data Curation, Writing - Review & Editing, Supervision.

## Statements and declarations

### Ethical considerations

The study was approved by Chulalongkorn University’s International Review Board (IRB) with the IRB No.0737/65. All procedures were conducted in accordance with the Declaration of Helsinki and applicable Thai and international regulations governing human research and privacy.

### Consent to participate

All participants provided written informed consent before enrollment.

### Consent for publication

Not applicable.

### Data Availability Statement

The data that support the findings of this study will be available from the corresponding author (MM) upon reasonable request. The dataset is not publicly available, as further analysis will be undertaken and for privacy and ethical reasons.

